# Oxidative and cardiovascular stress among professional divers in Egypt

**DOI:** 10.1101/2022.10.20.22281338

**Authors:** Hanie Salah, Ragaa M. El-Gazzar, Ekram W. Abd El-Wahab, Fahmy Charl

## Abstract

**Objectives:** Professional divers are exposed to unique multifactorial hazards in their working environment and cardiovascular effects are tremendous. Cardiovascular events are aggravated by diving-induced oxidative stress and account for one fourth of diving fatalities. The aim of this study was to assess the oxidative and cardiovascular stress in a group of professional divers in Alexandria, Egypt using a panel of biomarkers.

**Methods:** A comparative cross-sectional study was conducted between June 2017 and May 2018 at the General Naval Hospital in Alexandria. We recruited 50 professional divers (n=50) and a comparison group of 50 marine seafarers sharing similar maritime environments. Participants were clinically evaluated by electrocardiography (ECG) and assessment of some trace metals (Fe^+^, Cu^+^ and Zn^+^) and oxidative stress biomarkers (OSBMs; MDA, TAS, GST, GSH, GR, GPx, SOD and CAT). **Results**: Significant ECG abnormalities including short corrected QT interval, sinus bradycardia, left ventricular hypertrophy, early repolarization, 1^st^ degree heart block, intraventricular conduction defect were obvious among divers. The mean levels of FBG, LDH-C, Na^+^, Ca^+^, trace metals and OSBMs were significantly higher among professional divers compared to the marine seafarers (*p* < 0.5).

**Conclusion:** The risk of CVD in professional divers is alarmingly high and possibly contributed by OS. Including cardiovascular risk assessment in divers’ periodic examinations is of utmost importance.

## INTRODUCTION

Divers are employed in a wide range of activities, and all share unique hazards of their working environment ^1^. Diving exposes a body to an unnatural hyperbaric environment which can lead to increased gas dissolved in the tissues ^2^. The diving environment involves various stress factors including excess physical exercise demands, hemodynamics changes, thermal and mental stresses. Therefore, the disorders affecting divers are not limited to gas toxicity but can affect locomotion, respiration, circulation, kidney excretory and nervous systems ^3^.

Cardiovascular disease (CVD) has been identified as a possible contributing event to diving-related fatalities ^4–7^. Oxidative stress (OS) and chronic elevation of reactive oxygen species (ROS) is an additional contributing risk for cardiovascular disorders among divers ^8^. Indeed, when free radicals of nitrogen and oxygen species are increased, they damage the lipoproteins, DNA, lipids and proteins of cell structures. Thus, OS is mainly assessed by measuring oxidative stress biomarkers (OSBMs) which include malonyldialdehe (MDA), glutathione (GH), reduced glutathione (GSH), glutathione peroxidase (GPx), glutathione reductase (GRH), catalase enzyme, and superoxide dismutase (SOD) ^9^.

Likewise, trace metals are key players in the pathogenesis and evolution of in CVDs. Iron (Fe+), Copper (Cu+), and zinc (Zn+) are crucial for cardiovascular health and fitness. Their levels in the blood are disturbed in established CVDs ^10,11^. Trace elements are emerging novel tools for risk assessment of divers and its correlation with CVDs and OS ^11,12^.

The classical diver fitness and risk assessment in the pre-placement and periodic exam utilizes conventional tools stressing lipid risk factors and in some occasions cardiac stress factors without considering the early biomarkers of the increased risk among professional divers.

Accordingly, we will proceed in this study is to explore diving effects on some OSBMs and trace elements that may affect fitness and cardiovascular risk among professional divers compared to marine seafarers.

## METHODS

### Study setting, design and population

We conducted a comparative cross-sectional study between June 2017 and May 2018 at general naval hospital in Alexandria and authorized clinics for diving medical physicians. The target population comprised the national occupational professional male divers, fishermen divers, and freelance divers at different governmental and private companies and associations in Egypt.

Accordingly, we recruited a representative sample of eligible professional divers and a second group of non-diver marine workers matched in their socio-demographics and sharing the whole maritime environment but not exposed to diving hazards at their workplace. Professional divers and marines seek medical fitness examination at the general naval hospital in Alexandria as a perquisite for obtaining and/or renewal of their marine passport. For eligibility criteria, we included male divers in the age group 30-45 years, certified as professional divers with a diving license > 5 years, had accomplished at least 2 dives/month within decompression limits during the last year before examination, and having a valid complete signed logbook recording of: i) the diver medical fitness certificate (valid and singed by authorized diving medical doctor), ii) history of diving including the number of dives, maximum depth, maximum duration, diving bottom time, type of gas used in diving and diving plan. We excluded from the study divers and mariners with new overt CV symptoms, diabetes mellitus (DM), or any chronic inflammatory diseases such as rheumatoid arthritis, osteoartharosis, or any other musculoskeletal disorders.

### Sampling

Using G power 3.0.10, and based on a previous study on CVR factor assessment in professional divers ^13^, the minimum required sample size was 46 for each group. However, we ultimately enrolled 50 participants in each group. All eligible professional divers and seafarers fulfilling the inclusion criteria and accepted to participate in the study were consecutively enrolled until the required sample size was achieved.

### Clinical and laboratory assessments

All enrolled participants were interviewed for full occupational and medical history using a predesigned questionnaire form to collect background information on sociodemographics, lifestyle, physical activity and exercise, type of professional activity, number of professional and recreational dives during the last year; average and maximum depths and average and maximum durations, family and past medical history for relevant medical conditions to exclude co-morbidities.

The smoking index was calculated for current smokers by multiplying the number of cigarettes per day and the number of smoking years ^14^. Subjects were classified into either smoker [those who had smoked more than 100 cigarettes in their lifetime and currently still smoking or non-smoker [those who had not smoked more than 100 cigarettes and currently not smoking].

All divers were clinically evaluated by complete physical examinations (anthropometric measurements, complete general and systemic examination). Systemic clinical examination was done with a special emphasis on the cardiovascular system including resting heart rate, respiratory rate and blood pressure measurements that were done according to the standard procedures ^15^.

### Electrocardiography

A 12 lead surface electrocardiogram (ECG) was done for all patients in the supine position using a ECG device. The 12 lead ECG was recorded at a paper speed of 25 mm/s and 1 mV/cm standardization. The following data were noted: Rate/min, P–R interval (msec), QRS complex (msec), QT wave (msec), R–R minimum and R–R maximum which is the measurement of the long lead II during deep inspiration and the Sokolov of the precordial chest leads.

The corrected QT wave interval was calculated by the equation developed by Chenoweth et al.: The corrected QT = QT Interval / sqr (RR Interval) ^16–18^.

The main ECG criteria for left ventricular hypertrophy (LVH), including Sokolov-Lyon and Cornell Voltage indices, were calculated for all included individuals. The LVH was detected using the following formula: Sokolov-Lyon criteria= S_1/2_ +R_5/6_ > 35 ^19–21^

### Blood sampling

A five milliliter of venous blood was collected a aseptically from each participant in a vacutainer tube containing citrate or EDTA as anticoagulant. Tubes containing blood were centrifuged at 700 – 1000 *x*g for 10 minutes at 4**°**C. Top yellow plasma layer was pipetted off without disturbing the white buffy coat. The white buffy layer was then removed and discarded.

The RBCs were lysed in 4 times its volume of ice-cold HPLC grade water. The lysate was centrifuged at 100,000 *x*g for 15 minutes at 4**°**C. The collected plasma or erythrocyte lysate were stored on ice until assaying on the same day was done or frozen at −80 **°**C, thus samples can be left stable for at least one month.

### Biochemical tests

Biochemical analyses including fasting blood glucose (FBG), lipid profile [total cholesterol (TC), triglycerides (TG), high density lipoprotein cholesterol (HDL-C), low density lipoprotein cholesterol (LDL-C)], kidney function tests (blood urea, creatinine and uric acid), liver function tests (serum glutamic oxaloacetic transaminase (SGOT/ALT), serum glutamate-pyruvate transaminase (SGPT/AST) and serum bilirubin) and serum electrolytes (Na^+^, K^+^, Ca^+,^ Na^+^/K^+^ ratio) were done according to the standard laboratory procedures described by Poppiti and Sellers ^22^.

### Assessment of oxidative stress biomarkers

Maloney dialdehyde (MDA) ^23^, Superoxide dismutase enzyme (SOD) ^24^, Catalase enzyme (CAT) ^25^, Glutathione peroxidase (GPx) ^26^, Glutathione reductase (GHR) ^27^, Glutathione transferase (GST) ^28^, educed glutathione (GSH) ^29^, and total antioxidant status (TAS) ^30^ were assayed as previously described.

### Assessment of trace metals

Plasma trace metal levels of Fe^+^, Zn^+^ and Cu^+^ were analyzed by the colorimetric technique described by Kim and co-workers ^31^.

### Statistical data analysis

Data were fed to the computer and analyzed using IBM SPSS software package version 20.0 (Armonk, NY: IBM Corp). The Kolmogorov-Smirnov test was used to verify the normality of distribution of the different variables. Qualitative data were described using number and percent. Quantitative data was described using mean and standard deviation (SD). Chi-square test was used for categorical variables to compare between different groups. Fisher’s Exact test (FET) was used as a correction for chi-square when more than 20% of the cells have expected count less than 5. For normally distributed quantitative variables, *t*-test was used to compare between two studied groups. Pearson coefficient was used to correlate between two normally distributed quantitative variables. Mann Whitney U test was used for abnormally distributed quantitative variables to compare between the two studied groups. Wilcoxon signed ranks test was used for abnormally distributed quantitative variables to compare between two time periods. Significance of the obtained results was judged at the 5% level.

## RESULTS

### Sociodemographics and clinical data of the enrolled divers and seafarers

The mean age of divers was 36.48 ± 6.08 years, while that of the marine seafarers was (37.05 ± 5.16) years, (*p*= 0.615). The average employment duration for divers was comparable for that of the controls (13.10 ± 8.43 *vs* 13.0 ± 7.6 years, respectively).

Family history of CVD and/or DM was more frequently reported in the diver group than the seafarers although the differences between the two groups for both conditions were not statistically significant (*p* = 0.181 and *p* = 0.198 respectively).

A considerable number of the professional divers and marine seafarers were smokers, although the pattern of smoking did not differ significantly between the two studied groups (*p* >0.05).

The mean (±SD) BMI of the professional divers was 26.7 ± 3.7 with the majority being overweight. Type I obesity was prevailing among marine seafarers as evidenced by higher BMI, waist circumference and Waist\Height ratio compared to the professional divers (*p <0*.*05*). On the other hand, the divers showed a statistically significant higher mean SBP compared to the marines [131.18 ± 14.06 *vs* 124.3 ± 15.84 respectively, (*p*=0.02)] (Table 1).

**Table 1:** Sociodemographics and clinical data of the enrolled divers and seafarers

| Demographic and medical data | Divers<br>(n = 50) |  | Marines and<br>Seafarers<br>(n = 50) |  | Test of Sig. | p |
| --- | --- | --- | --- | --- | --- | --- |
|  | No. | % | No. | % |  |  |
| Age (years) |  |  |  |  |  |  |
| Mean ± SD | 36.48 ± 6.08 |  | 37.05 ± 5.16 |  | t=0.51 | 0.615 |
| Duration of employment ( years) | No. | % | No. | % |  |  |
| <5 | 10 | 20 | 6 | 12 | χ<sup>2</sup>=294 | 0.23 |
| 5 - <10 | 10 | 20 | 6 | 12 |  |  |
| ≥10 | 30 | 60 | 38 | 76 |  |  |
| Mean ± SD | 13.10 ± 8.43 |  | 13.0 ± 7.63 |  | U=1235 | 0.92 |
| Family history of cardiovascular diseases |  |  |  |  |  |  |
| No | 33 | 66 | 39 | 78 | χ<sup>2</sup>=1.79 | 0.181 |
| Yes | 17 | 34 | 11 | 22 |  |  |
| Family history of diabetes mellitus |  |  |  |  |  |  |
| No | 31 | 62 | 37 | 74 | χ<sup>2</sup>=1.65 | 0.198 |
| Yes | 19 | 38 | 13 | 26 |  |  |
| Smoking | 27 | 54 | 20 | 40 | χ<sup>2</sup>=1.97 | 0.161 |
| Smoking index (Mean ± SD) | 678.06± 255.64 |  | 735.47± 409.24 |  | U= 239.5 | 0.634 |
| Anthropometric measurements |  |  |  |  |  |  |
| BMI (kg/m<sup>2</sup>) |  |  |  |  |  |  |
| Normal (18.5–24.9) | 13 | 26 | 6 | 12 | χ<sup>2</sup>=8.20 | 0.017* |
| Overweight (25–29.9) | 26 | 52 | 20 | 40 |  |  |
| Obese (≤ 30) | 11 | 22 | 24 | 48 |  |  |
| Mean ± SD. | 26.66 ± 3.67 |  | 30.13 ± 4.38 |  | t=4.292* | <0.001* |
| Waist circumference (Mean ± SD) | 94.88 ± 11.59 |  | 101.9 ± 12.07 |  | t=2.975* | 0.004* |
| Waist \ height (Mean ± SD) | 0.53 ± 0.06 |  | 0.58 ± 0.07 |  | t=3.585* | 0.001* |
| Blood pressure (mmHg) |  |  |  |  |  |  |
| Normal (< 120/80 mmHg) | 11 | 22 | 15 | 30 | χ<sup>2</sup>=2.47 | 0.482 |
| Elevated (120-129/<80 mmHg) | 17 | 34 | 11 | 22 |  |  |
| High blood pressure 1 (130-139/80-90 mmHg) | 15 | 30 | 14 | 28 |  |  |
| High blood pressure 2 (>140/90 mmHg) | 7 | 14 | 10 | 20 |  |  |
| Systolic blood pressure (Mean ± SD) | 131.18 ± 14.06 |  | 124.3 ± 15.84 |  | t=2.30* | 0.024* |
| Diastolic blood pressure (Mean ± SD) | 77.70 ± 10.01 |  | 80.34 ± 9.61 |  | t=1.35 | 0.182 |

$\chi^2$ : Chi square test
FET: Fisher Exact test
MC: Monte Carlo
t: Student t-test
U: Mann Whitney test
p: p value for comparing between the studied groups; statistically significant at $p \leq 0.05$

### Biochemical changes among the study participants

The mean FBS level differed significantly among the divers and seafarers [89.0 ± 12.46 *vs* 100.5 ± 29.03 mg/dl, (*p*= 0.012)], though the levels were still within normal reference ranges (56-110 mg/dl). Likewise, renal function tests (blood urea, creatinine and uric acid levels) and liver function tests (SGOT, SGPT and serum bilirubin) were comparable between the two groups and within normal reference ranges (*p* >0.05).

Regarding the lipid profile, the mean plasma levels of TC, TG and LDL-C were comparable between the professional divers and seafarers (*p* > 0.05). On the other hand, the professional divers showed higher mean HDL-C levels compared to the seafarers [41.46 ± 4.01 *vs* 39.34 ± 4.34 respectively, (*p*= 0.013)] (Table 2).

**Table 2:** Biochemical changes and oxidative stress among the enrolled divers and marine seafarers

|  | Divers<br>(n = 50) | Marines and<br>Seafarers<br>(n = 50) | Test of Sig. | p |
| --- | --- | --- | --- | --- |
| **Oxidative stress biomarkers** |  |  |  |  |
| MDA (0.67-2.44 $\mu\text{m}/\text{l}$) | | | | |
| Min. – Max. | 4.90 – 22.0 | 2.50 – 15.0 |  |  |
| Mean ± SD. | 13.54 ± 4.08 | 6.31 ± 2.66 | U= 183.50* | <0.001* |
| TAS (0.5-2 mmol/dl) |  |  |  |  |
| Min. – Max. | 0.29 – 2.0 | 0.50 – 2.30 |  |  |
| Mean ± SD. | 0.84 ± 0.43 | 1.40 ± 0.45 | U= 444.50* | <0.001* |
| GST (3.38 $\mu\text{m}/\text{dl}$) | | | | |
| Min. – Max. | 0.50 – 6.20 | 2.10 – 12.30 |  |  |
| Mean ± SD. | 2.42 ± 1.29 | 6.43 ± 2.40 | U= 153.50* | <0.001* |
| GSH (24-36 IU/ml) |  |  |  |  |
| Range | 8.50 – 27.50 | 16.40 – 33.0 |  |  |
| Mean ± SD. | 15.22 ± 4.15 | 24.34 ± 3.37 | U= 145.00* | <0.001* |
| GR (IU/ml) |  |  |  |  |
| Min. – Max. | 5.90 – 25.20 | 14.30 – 40.0 |  |  |
| Mean ± SD. | 11.66 ± 4.70 | 24.08 ± 5.16 | U= 104.50* | <0.001* |
| GPx (up-8 U/ml) |  |  |  |  |
| Min. – Max. | 0.01 – 0.35 | 0.05 – 0.51 |  |  |
| Mean ± SD. | 0.11 ± 0.09 | 0.24 ± 0.12 | U= 406.00* | <0.001* |
| SOD (100 U/ml) |  |  |  |  |
| Min. – Max. | 98.0 – 211.0 | 144.0 – 345.0 |  |  |
| Mean ± SD. | 140.5 ± 27.59 | 243.2 ± 40.19 | t= 14.894* | <0.001* |
| CAT (IU/ml) |  |  |  |  |
| Range | 214.0 – 440.0 | 265.0 – 562.0 |  |  |
| Mean ± SD. | 298.7 ± 58.20 | 436.2 ± 76.23 | t= 10.136* | <0.001* |
| **Trace metal** |  |  |  |  |
| Fe<sup>+</sup> (65-180 $\mu\text{m}/\text{dl}$). | | | | |
| Min. – Max. | 65.0 – 251.0 | 25.0 – 181.0 |  |  |
| Mean ± SD. | 147.6 ± 38.08 | 101.1 ± 38.17 | t= 6.101* | <0.001* |
| Cu<sup>+</sup> (70-150 $\mu\text{m}/\text{dl}$). | | | | |
| Min. – Max. | 132.0 – 449.0 | 53.0 – 169.0 |  |  |
| Mean ± SD. | 271.3 ± 75.01 | 100.8 ± 30.20 | t= 14.908* | <0.001* |
| Zn<sup>+</sup> (70-100 $\mu\text{m}/\text{dl}$). | | | | |
| Min. – Max. | 40.0 – 189.0 | 75.0 – 163.0 |  |  |
| Mean ± SD. | 85.52 ± 27.37 | 116.6 ± 21.95 | t= 6.264* | <0.001* |
| **Electrolytes** |  |  |  |  |
| Ca<sup>+</sup> (9-11 mmol/L) |  |  |  |  |
| Mean ± SD. | 9.44±0.52 | 9.19±0.60 | t= 2.197* | 0.030* |
| K<sup>+</sup> (3.5-5.2 mmol/L) |  |  |  |  |
| Mean ± SD. | 4.20 ± 0.39 | 4.26 ± 0.34 | t= 0.936 | 0.352 |
| Na<sup>+</sup> (135-145 mmol/L) |  |  |  |  |
| Mean ± SD. | 141.72±3.53 | 143.26±3.99 | t= 2.043* | 0.044* |
| Na<sup>+</sup>/K<sup>+</sup> ratio |  |  |  |  |
| Mean ± SD. | 34.04 ± 3.08 | 33.82 ± 3.06 | t= 0.359 | 0.72 |
| **Metabolic tests** |  |  |  |  |
| **FBS (56-110 mg %)** |  |  |  |  |
| Mean ± SD. | 89.0 ± 12.46 | 100.5 ± 29.03 | t= 2.574* | 0.012* |
| Urea (20-50 ml / dl) |  |  |  |  |
| Min. – Max. | 23.0 – 36.0 | 21.0 – 42.0 |  |  |
| Mean ± SD. | 28.74 ± 3.49 | 30.28 ± 5.38 | t= 1.698 | 0.093 |
| Creatinine (0.5-1.5 ml /dl) |  |  |  |  |
| Min. – Max. | 0.61 – 1.0 | 0.60 – 1.10 |  |  |
| Mean ± SD. | 0.78 ± 0.10 | 0.80 ± 0.12 | t= 0.587 | 0.559 |
| Uric Acid (2.5-6 ml /dl) |  |  |  |  |
| Min. – Max. | 3.0 – 7.0 | 3.0 – 7.0 |  |  |
| Mean ± SD. | 4.94 ± 1.02 | 5.0 ± 0.90 | t= 0.312 | 0.756 |
| SGOT (up to 40 ml /dl) |  |  |  |  |
| Min. – Max. | 13.0 – 35.0 | 12.0 – 38.0 |  |  |
| Mean ± SD. | 23.72 ± 6.76 | 25.06 ± 7.54 | t= 0.935 | 0.352 |
| SGPT (up to 40 ml /dl) |  |  |  |  |
| Min. – Max. | 13.0 – 39.0 | 11.0 – 56.0 |  |  |
| Mean ± SD. | 22.60 ± 5.80 | 24.66 ± 8.77 | t= 1.386 | 0.169 |
| Bilirubin (0.5-1.5 ml /dl) |  |  |  |  |
| Min. – Max. | 0.46 – 1.05 | 0.44 – 1.0 |  |  |
| Mean ± SD. | 0.68 ± 0.13 | 0.72 ± 0.12 | t= 1.916 | 0.058 |
| **Lipid profile** |  |  |  |  |
| Triglycerides (50-150 mg /dl) |  |  |  |  |
| Mean ± SD. | 115.3 ± 26.41 | 119.2 ± 26.54 | t= 0.748 | 0.456 |
| Total cholesterol (125-200 mg /dl) |  |  |  |  |
| Mean ± SD. | 182.3 ± 19.66 | 177.5 ± 20.84 | t= 1.19 | 0.237 |
| HDL-C (40-80 mg/dl) |  |  |  |  |
| Mean ± SD. | 41.46 ± 4.01 | 39.34 ± 4.34 | t= 2.538* | 0.013* |
| LDL-C (85-125 mg/dl) |  |  |  |  |
| Mean ± SD. | 117.6 ± 17.31 | 114.3 ± 17.79 | t= 0.952 | 0.344 |

t: Student *t*-test
U: Mann Whitney test
p: *p* value for comparing between the studied groups; statistically significant at $p \leq 0.05$

### Biomarkers of oxidative stress

Regarding lipid peroxidation, the mean MDA level was high in both groups compared to its reference range (0.67-2.44 µml), although it was significantly higher among the professional divers compared to the seafarers [13.54 ± 4.08 vs 6.31 ± 2.66 µml, (*p* <0.001)].

As for the antioxidant system, the TAS did not increased in both groups. Its mean level remained within the normal reference range (0.5–2.0 µml), although it was significantly lower in the divers than in the marine seafarers [0.84 ± 0.43 *vs* 1.40 ± 0.45 µml respectively, (*p* <0.001)]. The ean CAT, GPx, GSH, GR, and GST enzyme levels were significantly reduced in the professional divers (*p* <0.001). On the other hand, the mean SOD level was high in both groups compared to the reference range (100 U/ml), although it was significantly higher among controls compared to the professional divers [140.5 ± 27.59 *vs* 243.2 ± 40.19 U/ml, (*p* <0.001)] (Table 2).

### Trace metals

The mean Fe^+^ level was significantly lower among the professional divers than in the controls [101.1 ± 38.17 *vs* 147.6 ± 38.08 µm/dl, (*p* <0.001)]. Similarly, the mean level of Zn^+^ was significantly lower among the professional divers than in the seafarers [85.52 ± 27.37 *vs* 116.6 ± 21.95 µm/dl, (*p* <0.001)]. Conversely, the professional divers showed higher mean level of Cu^+^ compared to the seafarers [271.3 ± 75.01 *vs* 100.8 ± 30.20 µm/dl, (*p* <0.001)] (Table 2).

### Electrolyte profile

The mean plasma Na^+^, K^+^, and Ca^+^, levels were comparable between the divers and the seafarers (9.44 ± 0.52, 4.20 ± 0.39 and 141.72 ± 3.53 *vs* 9.19 ± 0.60, 4.26 ± 0.34 and 143.26 ± 3.99 mmol/L respectively) although the differences for Ca^+^ and Na^+^ between the two groups were statistically significant (*p* = 0.030 and *p* = 0.044 respectively). On the other hand, the mean Na^+^/K^+^ ratio did not differ significantly between the two groups [34.04 ± 3.08 *vs* 33.82 ± 3.06 mmol/L, (*p* = 0.720)] (Table 2).

### Quantitative ECG changes in the enrolled professional divers and seafarers

Abnormal ECG changes were found in 22 divers (44.0%) and 7 seafarers (14.0%). The mean heart rate (± SD) was significantly slower among the professional divers compared to the seafarers (68.90 ± 16.06 *vs* 78.16 ± 15.42 beat/min respectively, *p* = 0.004), although both values were still within the normal range. The professional divers showed more prolonged mean (±SD) P-R interval [168.8 ± 28.62 *vs* 140.8 ± 21.37 msec respectively, (*p* < 0.001)] and QRS [85.20 ± 20.92 *vs* 75.60 ± 17.75 msec respectively, (*p* = 0.002)] compared to the seafarers. There was no significant difference between the two groups regarding the mean QT segment (*p* = 0.658), although the mean (±SD) corrected QT segment differed significantly among the professional divers and the seafarers [392.1 ± 22.67 *vs* 412.5 ± 37.76 msec respectively, (*p*= 0.002)]. The mean Sokolov–Lyon score (S_1/2_+R_5/6_) was significantly higher in the professional divers (22.68 ± 6.18 mV) than in the controls (12.50 ± 4.09 mV) (*p* <0.001). Likewise, the R-R interval, the R-R, the R-R maximum and the mean R-R, all significantly differed among both groups (*p* < 0.001) (Table 3).

**Table 3:**
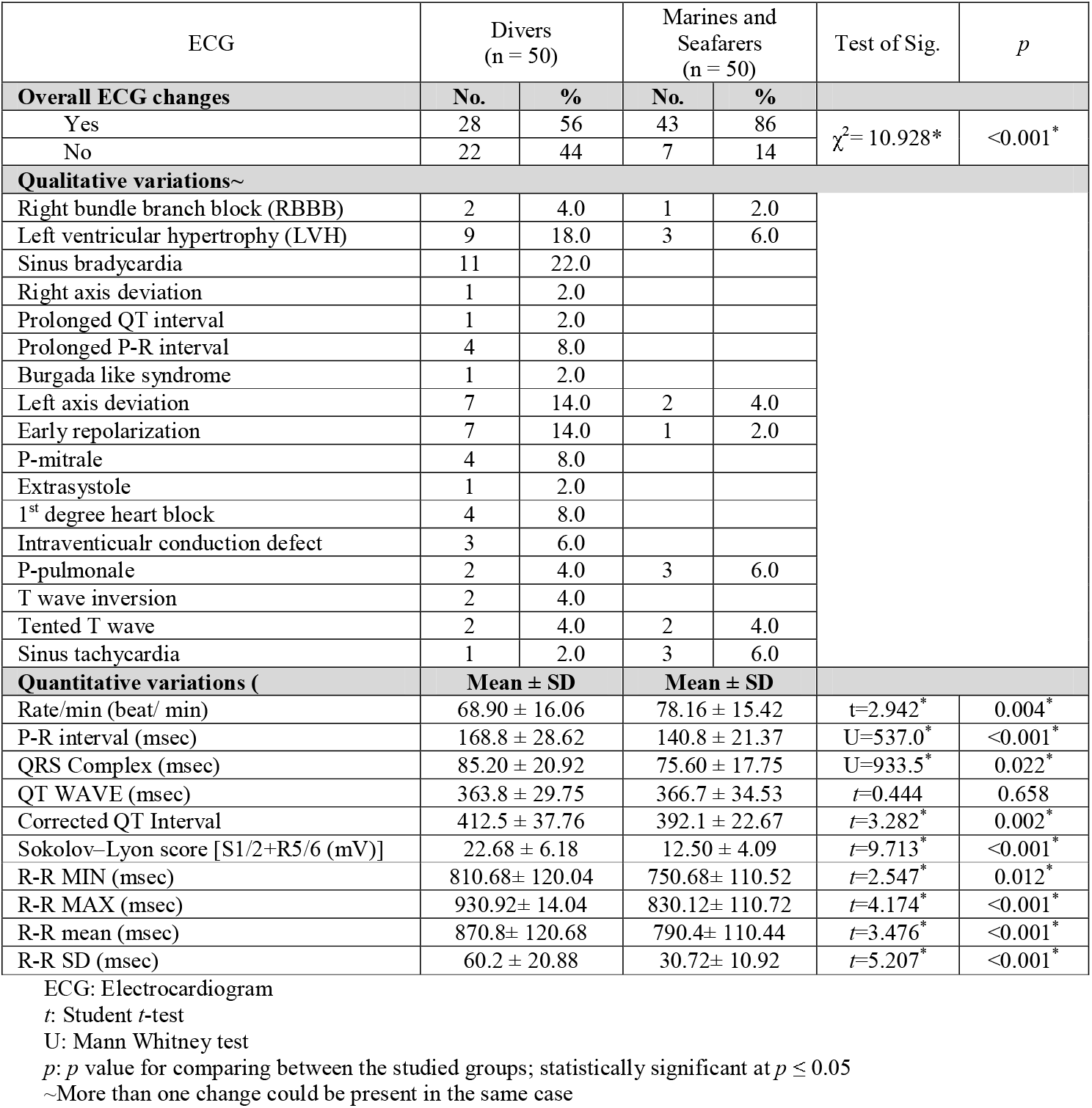
Quantitative and qualitative electrocardiogram changes among the enrolled divers and marine seafarers

### Qualitative ECG changes in the enrolled professional divers and seafarers

The most common variations among the professional divers were sinus bradycardia (22.0%), LVH (18.0%), left axis deviations (14.0%), early repolarization (14.0%), athletes heart P-mitrale (8.0%), first degree heart block (8.0%), intraventricular conduction defect (6.0%), Right bundle branch block (RBBB) (4.0%), P-Pulmonale (4.0%), T wave inversion (4.0%), right axis deviation (2.0%), prolonged QT (2.0%), Burgada like syndrome (2.0%), peaked T wave (2.0%), and sinus tachycardia (2.0%). The overall variations in the studied divers were more common in both number and context compared to the marine seafarers. The latter mainly showed sinus tachycardia (6.0%), LVH (6.0%), P-pulmonale (6.0%), left axis deviation (4.0%), tented T-wave (4.0%), RBBB (4.0%) and early repolarization (2.0%) (Table 3).

## DISCUSSION

In the present study, we used a multimarkers approch to assess the cardiovascular and OS in professional divers. Our panel of biomarkers included some OSBMs, trace metals, electrolytes and electrophysiologic changes in the ECG.

The study population of professional diversa and seafarers were found matched as regards their age, duration of emplyment as well as the history of smoking and their family history of CVDs and DM. Overweight and obesity were common features among searfarers although the percentage was also high among divers. This was also reported in a study done in France on 200 professional divers where 43.6% of divers were overweight or obese ^21^. Also, in a stduy measuring the BMI in 1115 german and Itlalain sailors, approximtely 40% of evaluated subjects were overweight, and more than the 10% of them were obese ^32^. The relative differencesin BMI could reflect the increased physical activites done by professional divers during their tasks and different lifestyles led by seafares. Increased BMI was was previously reported to be associated with an increase in the 10 years and lifetime risks of CVD ^33–35^. The mean WHtR was significantly lower in divers than in controls and correlated with prolonged corrected QT interval which reflects disturbance in the electrophysiologic status. This is agreed with the previous studies that reported WHtR as a predictor of CVD risk ^35–39^.

The BP was obviously elevated in most participants albeit with a higher percentage among divers. This came in agreement with other studies where the percentage of high BP among divers varied between 6.5% and 64.0% ^21,40^. Increased prevalence of high SBP was also reported among seafarers in Denmark (44.7%) ^41^ and Germany (33.8%) ^42^. This may be due to the higher OS caused by diving.

Apart from HDL-C, lipid profile measurments did not differ significantly between the two study groups where levels remained within normal ranges. This contradicts reports in other studies that TC was raised in 50% of professional divers ^21^. Another study reported higher TG levels among 41.6% of seafarers ^43^.

Liver and kidney function tests together with uric acid were within normal ranges among the two studied groups. This means that there were no kidney or liver conditions that could influence the other assessed laboratory parameter particularly OSBMs and trace metals which can refelct liver and kidney diseases ^44,45^. Likewise, normal mean FBS but significantly lower among divers than seafarers could reflect excess physical efforts done by divers during their tasks and to a less extent to the diving environment as as postulated in similar studies ^46,47^.

The ECG is necessary for early diagnosis of cardiovascular pathology. Few studies addressed the qualitative changes of the ECG. In the present study, the professional divers showed 5 times higher Odds of having electrophysiological changes compared to marine seafarers. Sinus bradycardia, LVH, early repolarization, 1^st^ degree heart block, intraventicular conduction defect were obvious changes. Indeed, the high ambient pressure and increased gas densities can lead to cardiac decompensation. Sinus arrhythmia is frequent in athletes as a result of strenuous exercise wherease other changes are considered to be pathological ^48^. In a study done on 225 randomly selected professional divers, significant ECG abnormalities such as incomplete IRBBB, right QRS axis deviation, sinus tachycardia, early repolarization, ventricular extrasystoles, ST elevation and sinus arrhythmia and T-wave inversion were reported ^49^. In fact, electrophysiological changes are important clues as they are more likely to precipitating or aggravate arrhythmias in divers and hence more relevant etiological factor for cardiac related deaths rather than ischemic coronary vascular changes ^50–53^. Adding electrophysiological changes to the CVR assessment will help also in risk stratification. It is worth noting that no other studies described corrected QT wave except a single recent study done by Zizaziabari et al., in 2019. They reported significant change in the pulse rate, corrected QT, and T wave before and after diving but did not define a cutoff point ^54^. ECG changes were also found to be a more specific biomarkers as OS changes could occur in other physiological and pathological conditions ^20^. ECG changes as a biomarker are characterized by large availability, simple interpretation, and cost-effectiveness and should therefore be preferred in the CVD risk assessment ^20^.

Evidence for increased OS was found in both studied groups as manifested by abnormal levels of OSBMs. This was more obvious among divers compared to seafarers and came in agreement with many studies which spotted increased OS to be associated with increased cardiovascular risks and hypertension ^55–58^. Other studies showed that the assessed CVR was higher the higher the OS levels was ^59^. Meanwhile, the current results support the postulation that diving and excercises increase the OS in divers ^60,61^. Remarkably, MDA, SOD, GST and TAS were more sensitive in reflecting OS than other biomarkers where they demonstrated the highest percent of changes compared to the normal references values.

The role of trace metals and OS in the pathogesis of CVDs is well established ^62–64^. Also, OS is linked to increased levels of Cu^+^ and decreased levels of Fe^+^ and Zn^+ 64–66^. This was in concordance with the present work, where the levels of Fe^+^ and Zn^+^ were significantly lower in divers compared to seafares whereas the level of Cu^+^ was significantly higher. Few research mentioned the effect of diving on trace metals especilly the staturation deep diving which affects the hemoglobin iron level ^67^.

In the present study, the mean serum electrolyte (Ca^+^, Na^+^ and K^+^) levels among the two groups were within the normal references values, although higher level of Ca^+^ and lower level of Na^+^ were seen in the professtional divers compared to the seafareres. A recent study showed that high blood Ca^+^ level may increases arterial wall stiffness and subsquently the 10 years CVD risk as assessed by the Farmingham score ^68^. This might represent another novel factor which can lead to increase in the CVD risk. Indeed, hyponatrimea may be attributed to the repeated physical activity done by divers during their work and the physical effect of diving in agreement with statements of Luhker and Rosner ^69,70^.

In conclusion, cardiovascular stress in professional divers is alarmingly high and OS as well as trace metal imbalance are possible contributing factors.

The electorphsyiological changes of the cardiac activties detected by ECG are powerful CVD biomarkers. Furthermore, OS and trace metal levels are linked to the pathophysiology of CVDs and together with electorphsyiological changes of ECG could be validated as biomarkers for CVR assessment.

## Data Availability

All data produced in the present work are contained in the manuscript

## Ethical considerations

### Ethical approval

The study was approved by the institutional review board and the Ethics Committee of the High Institute of Public Health-Alexandria University. The research was conducted in accordance with the ethical guidelines of Helsinki’s Declaration (2013). Data sheets were coded with numbers to maintain the anonymity and confidentiality of patient’s data.

This article does not contain any studies with animals performed by any of the authors.

### Informed consent

All participants signed an informed written consent after explaining the aim and concerns of the study.

### Funding

None

## References

1. Mitchell S, Bennett M. Clearance to dive and fitness for work. In: Neuman T, Thom S, editors. The physiology and medicine of hyperbaric oxygen therapy. Philadelphia, PA: Saunders; 2008. p. 65–94.

2. Pendergast DR, Lundgren CE. The physiology and pathophysiology of the hyperbaric and diving environments. J Appl Physiol (1985). 2009 Jan;106(1):274–5.

3. Levett DZ, Millar IL. Bubble trouble: a review of diving physiology and disease. Postgrad Med J. 2008 Nov;84(997):571–8.

4. Denoble PJ, Caruso JL, Dear Gde L, Pieper CF, Vann RD. Common causes of open-circuit recreational diving fatalities. Undersea Hyperb Med. 2008 Nov-Dec;35(6):393–406.

5. Mitchell SJ, Bove AA. Medical screening of recreational divers for cardiovascular disease: consensus discussion at the Divers Alert Network Fatality Workshop. Undersea Hyperb Med. 2011 Jul-Aug;38(4):289–96.

6. Bove AA. The cardiovascular system and diving risk. Undersea Hyperb Med. 2011 Jul-Aug;38(4):261–9.

7. Edmonds C, Caruso J. Diving fatality investigations: recent changes. Diving Hyperb Med. 2014 Jun;44(2):91–6.

8. Tirapelli CR. Oxidative Stress and Vascular Disease. Curr Hypertens Rev. 2020;16(3):162.

9. Ceconi C, Boraso A, Cargnoni A, Ferrari R. Oxidative stress in cardiovascular disease: myth or fact? Archives of biochemistry and biophysics. 2003;420(2):217–21.

10. Shokrzadeh M, Ghaemian A, Salehifar E, Aliakbari S, Saravi SSS, Ebrahimi P. Serum zinc and copper levels in ischemic cardiomyopathy. Biological trace element research. 2009;127(2):116.

11. Valko M, Morris H, Cronin M. Metals, toxicity and oxidative stress. Current medicinal chemistry. 2005;12(10):1161–208.

12. Valko M, Leibfritz D, Moncol J, Cronin MT, Mazur M, Telser J. Free radicals and antioxidants in normal physiological functions and human disease. The international journal of biochemistry & cell biology. 2007;39(1):44–84.

13. Pougnet R, Costanzo LD, Lodde B, Henckes A, Dherbecourt L, Lucas D, et al. Cardiovascular risk factors and cardiovascular risk assessment in professional divers. Int Marit Health. 2012;63(3):164–9.

14. Centers for Disease Control Prevention (CDC). Current cigarette smoking among adults in the United States. USA: CDC; 2016.

15. Colantonio LD, Booth III JN, Bress AP, Whelton PK, Shimbo D, Levitan EB, et al. 2017 ACC/AHA blood pressure treatment guideline recommendations and cardiovascular risk. Journal of the American College of Cardiology. 2018;72(11):1187–97.

16. Bazett HC. An analysis of the time relations of electrocardiograms. Heart. 1920;7:353–70.

17. Israel SA, Irvine JM, Cheng A, Wiederhold MD, Wiederhold BK. ECG to identify individuals. Pattern recognition. 2005;38(1):133–42.

18. Chenoweth JA, Hougham AM, Colby DK, Ford JB, Sandhu J, Albertson TE, et al. Monitoring the corrected QT in the acute care setting: A comparison of the 12-lead ECG and bedside monitor. The American journal of emergency medicine. 2018;36(5):777–9.

19. Tamama K, Kanda T, Osada M, Nagai R, Suzuki T, Kobayashi I. Detection of left ventricular enlargement by electrocariography. Journal of medicine. 1998;29(3-4):231–6.

20. Tocci G, Figliuzzi I, Presta V, El Halabieh NA, Citoni B, Coluccia R, et al. Adding markers of organ damage to risk score models improves cardiovascular risk assessment: Prospective analysis of a large cohort of adult outpatients. International journal of cardiology. 2017;248:342–8.

21. Pougnet R, Di Costanzo L, Loddé B, Henckes A, Dherbecourt L, Lucas D, et al. Cardiovascular risk factors and cardiovascular risk assessment in professional divers. International maritime health. 2012;63(3):164–9.

22. Poppiti JA, Sellers C. Practical techniques for laboratory analysis: CRC Press; 1994.

23. Ishizuka I. Chemistry and functional distribution of sulfoglycolipids. Progress in lipid research. 1997;36(4):245–319.

24. Arafa MH, Mohammad NS, Atteia HH, Abd-Elaziz HR. Protective effect of resveratrol against doxorubicin-induced cardiac toxicity and fibrosis in male experimental rats. Journal of physiology and biochemistry. 2014;70(3):701–11.

25. Aebi H. Catalase in vitro Methods Enzymol 105:121–126. Find this article online. 1984.

26. Paglia DE, Valentine WN. Studies on the quantitative and qualitative characterization of erythrocyte glutathione peroxidase. The Journal of laboratory and clinical medicine. 1967;70(1):158–69.

27. Goldberg D, Spooner R. Methods of enzymatic analysis. Bergmeyer HV. 1983;3:p258–65.

28. Habig WH, Pabst MJ, Jakoby WB. Glutathione S-transferases the first enzymatic step in mercapturic acid formation. Journal of biological Chemistry. 1974;249(22):7130–9.

29. Thomas EL, Grisham MB, Jefferson MM. [41] Cytotoxicity of chloramines. Methods in enzymology: Elsevier; 1986. p. 585–93.

30. Koracevic D, Koracevic G, Djordjevic V, Andrejevic S, Cosic V. Method for the measurement of antioxidant activity in human fluids. Journal of clinical pathology. 2001;54(5):356–61.

31. Kim KB, Kim H, Song EJ, Kim S, Noh I, Kim C. A cap-type Schiff base acting as a fluorescence sensor for zinc (II) and a colorimetric sensor for iron (II), copper (II), and zinc (II) in aqueous media. Dalton Transactions. 2013;42(47):16569–77.

32. Nittari G, Tomassoni D, Di Canio M, Traini E, Pirillo I, Minciacchi A, et al. Overweight among seafarers working on board merchant ships. BMC public health. 2019;19(1):45.

33. Khan SS, Ning H, Wilkins JT, Allen N, Carnethon M, Berry JD, et al. Association of body mass index with lifetime risk of cardiovascular disease and compression of morbidity. JAMA cardiology. 2018;3(4):280–7.

34. Lamon-Fava S, Wilson PW, Schaefer EJ. Impact of body mass index on coronary heart disease risk factors in men and women: the Framingham Offspring Study. Arteriosclerosis, thrombosis, and vascular biology. 1996;16(12):1509–15.

35. Melander O, Newton-Cheh C, Almgren P, Hedblad B, Berglund G, Engström G, et al. Novel and conventional biomarkers for prediction of incident cardiovascular events in the community. Jama. 2009;302(1):49–57.

36. Cai L, Liu A, Zhang Y, Wang P. Waist-to-height ratio and cardiovascular risk factors among Chinese adults in Beijing. PloS one. 2013;8(7):e69298.

37. Garg N, Muduli SK, Kapoor A, Tewari S, Kumar S, Khanna R, et al. Comparison of different cardiovascular risk score calculators for cardiovascular risk prediction and guideline recommended statin uses. Indian heart journal. 2017;69(4):458–63.

38. Sabah KMN, Chowdhury AW, Khan HLR, Hasan AH, Haque S, Ali S, et al. Body mass index and waist/height ratio for prediction of severity of coronary artery disease. BMC research notes. 2014;7(1):246.

39. Madden C, Putukian M, McCarty E, Young C. Netter’s Sports Medicine E-Book: Elsevier Health Sciences; 2013.

40. Åsmul K, Irgens Å, Grønning M, Møllerløkken A. Diving and long-term cardiovascular health. Occupational Medicine. 2017;67(5):371–6.

41. Tu M, Jepsen JR. Hypertension among Danish seafarers. International maritime health. 2016;67(4):196–204.

42. Oldenburg M, Jensen H-J, Latza U, Baur X. Coronary risks among seafarers aboard German-flagged ships. International archives of occupational and environmental health. 2008;81(6):735–41.

43. Oldenburg M. Risk of cardiovascular diseases in seafarers. International maritime health. 2014;65(2):53–7.

44. Romero FJ, Bosch-Morell F, Romero MJ, Jareño EJ, Romero B, Marín N, et al. Lipid peroxidation products and antioxidants in human disease. Environmental health perspectives. 1998;106(suppl 5):1229–34.

45. Halliwell B, Cross CE. Oxygen-derived species: their relation to human disease and environmental stress. Environmental health perspectives. 1994;102(suppl 10):5–12.

46. Dear G, Pollock N, Uguccioni D, Dovenbarger J, Feinglos M, Moon R. Plasma glucose responses in recreational divers with insulin-requiring diabetes. 2004.

47. Sponsiello N, Cialoni D, Pieri M, Marroni A. Cellular Glucose Uptake During Breath-Hold Diving in Experienced Male Breath-Hold Divers. Sports medicine-open. 2018;4(1):14.

48. Kiss O, Sydo N, Vargha P, Edes E, Merkely G, Sydo T, et al. Prevalence of physiological and pathological electrocardiographic findings in Hungarian athletes. Acta Physiol Hung. 2015 Jun;102(2):228–37.

49. Gunes AE, Cimsit M. The prevalence of electrocardiogram abnormalities in professional divers. Diving and Hyperbaric Medicine. 2017;47(1):55–8.

50. Greenland P, Alpert JS, Beller GA, Benjamin EJ, Budoff MJ, Fayad ZA, et al. 2010 ACCF/AHA guideline for assessment of cardiovascular risk in asymptomatic adults: a report of the American College of Cardiology Foundation/American Heart Association task force on practice guidelines developed in collaboration with the American Society of Echocardiography, American Society of Nuclear Cardiology, Society of Atherosclerosis Imaging and Prevention, Society for Cardiovascular Angiography and Interventions, Society of Cardiovascular Computed Tomography, and Society for Cardiovascular Magnetic Resonance. Journal of the American College of Cardiology. 2010;56(25):e50–e103.

51. Boässon M, Rienks R. Arrhythmogenicity of scuba diving: Holter monitoring in a hyperbaric environment. Undersea & hyperbaric medicine: journal of the Undersea and Hyperbaric Medical Society, Inc. 2019;46(4):421–7.

52. Shenasa M, Shenasa H. Hypertension, left ventricular hypertrophy, and sudden cardiac death. International journal of cardiology. 2017;237:60–3.

53. Denoble PJ. Hypertension, Left Ventricular Hypertrophy and Sudden Cardiac Death in Scuba Diving. Wound Care & Hyperbaric Medicine. 2013;4(3):21–6.

54. ZiaZiabari SM, Asadi P, Shakiba M, Habibi A, Shokrollahi A, Seidshekan J, et al. Electrocardiographic Changes in Scuba Divers: A Quasi-Experimental Study in Iran. 2019. [Scuba diving, Electrocardiography, Heart rate, Iran.]. 2019 2019-09-01;4(4):4–6.

55. Bastani A, Rajabi S, Daliran A, Saadat H, Karimi-Busheri F. Oxidant and antioxidant status in coronary artery disease. Biomedical reports. 2018;9(4):327–32.

56. Ho E, Galougahi KK, Liu C-C, Bhindi R, Figtree GA. Biological markers of oxidative stress: applications to cardiovascular research and practice. Redox biology. 2013;1(1):483–91.

57. Frijhoff J, Winyard PG, Zarkovic N, Davies SS, Stocker R, Cheng D, et al. Clinical relevance of biomarkers of oxidative stress. Antioxidants & redox signaling. 2015;23(14):1144–70.

58. Birben E, Sahiner UM, Sackesen C, Erzurum S, Kalayci O. Oxidative stress and antioxidant defense. World Allergy Organ J. 2012 Jan;5(1):9–19.

59. Poredoš P, Ježovnik MK. Markers of preclinical atherosclerosis and their clinical relevance. Vasa. 2015;44(4):247–56.

60. Perović A, Unić A, Dumić J. Recreational scuba diving: negative or positive effects of oxidative and cardiovascular stress? Biochemia medica: Biochemia medica. 2014;24(2):235–47.

61. Gorman D, Sames C, Mitchell S. Routine occupational dive medical examinations. 2009.

62. Momtaz M, Mughal N, Siddique A, Mahboob T. CHANGES IN BLOOD LEVELS OF TRACE ELEMENTS AND ELECTROLYTES IN HYP ERTENSIVE PATIENTS. Medical Journal of The Islamic Republic of Iran (MJIRI). 2000;14(2):115–8.

63. Nagarajrao R. Study of trace elements and malondialdehyde levels in cardiovascular disease patients. Int J Adv Res Biol Sci. 2014;1(9):25–32.

64. Osredkar J, Sustar N. Copper and zinc, biological role and significance of copper/zinc imbalance. J Clinic Toxicol S. 2011;3(2161):0495.

65. Marreiro D, Cruz K, Morais J, Beserra J, Severo J, de Oliveira A. Zinc and oxidative stress: current mechanisms. Antioxidants. 2017;6(2):24.

66. Sarkar PD, Ramprasad N, Sarkar ID, Shivaprakash T. Study of oxidative stress and trace element levels in patients with alcoholic and non-alcoholic coronary artery disease. Indian journal of physiology and pharmacology. 2007;51(2):141.

67. Nakabayashi K, Mizukami H, Hashimoto A, Oiwa H. CHANGE IN RED BLOOD CELL PRODUCTION RATE DURING A 330 MSW SATURATION DIVE SIMULATION. 1991.

68. Park B, Lee YJ. Borderline high serum calcium levels are associated with arterial stiffness and 10Dyear cardiovascular disease risk determined by Framingham risk score. The Journal of Clinical Hypertension. 2019;21(5):668–73.

69. Rosner MH, Kirven J. Exercise-associated hyponatremia. Clinical Journal of the American Society of Nephrology. 2007;2(1):151–61.

70. Lühker O, Berger MM, Pohlmann A, Hotz L, Gruhlke T, Hochreiter M. Changes in acid–base and ion balance during exercise in normoxia and normobaric hypoxia. European journal of applied physiology. 2017;117(11):2251–61.

